# ‘Trained immunity’ from *Mycobacterium* spp. (environmental or BCG) exposure predicts protection from Coronavirus disease 2019 (COVID-19)

**DOI:** 10.1101/2021.02.11.20233593

**Authors:** Samer Singh, Dhiraj Kishore, Rakesh K. Singh

## Abstract

Endeavors to identify potentially protective variables for COVID-19 impact on certain populations have remained a priority. Multiple attempts have been made to attribute the reduced COVID-19 impact on populations to their bacillus Calmette–Guérin (BCG) vaccination coverage ignoring the fact that the effect of childhood BCG vaccination wanes within 5 years while most of the COVID-19 cases and deaths have occurred in aged with comorbidities. Since the supposed protection being investigated could come from heterologous ‘trained immunity’ (TI) conferred by exposure to *Mycobacterium* spp. (*i*.*e*., environmental and BCG), it is argued that the estimates of the prevalence of TI of populations currently available as latent tuberculosis infection (LTBI) prevalence would be a better variable to evaluate such assertions. Indeed, when we analyze the European populations (twenty-four), and erstwhile East and West Germany populations completely disregarding their BCG vaccination coverage, the populations with higher TI prevalence consistently display reduced COVID-19 impact as compared to their lower TI prevalence neighbors. The TI estimates of the populations not the BCG coverage *per se*, negatively correlated with pandemic phase-matched COVID-19 incidences (*r*(24): - 0.79 to -0.57; p-value: <0.004), mortality (*r*(24): -0.63 to -0.45; *p*-value: <0.03), and interim case fatality rates(i-CFR) data. To decisively arrive at dependable conclusions about the potential protective benefit gained from BCG vaccination in COVID-19, the ongoing/planned randomized controlled trials should consciously consider including measures of TI as - a) all individuals immunized do not respond equally, b)small study groups from higher background TI could fail to indicate any protective effect.

Summary Box

What is already known?

- Previously, COVID-19 incidence (SARS-CoV-2 infections) and mortality datasets of disparate populations with regard to phase of pandemic, demographics, medical infrastructure etc. have been modelled to negatively associate with highly transformed BCG vaccination coverage and policy of the countries.
- Recently BCG vaccination has been linked to risk of COVID-19 using vaccinated individuals from disparate populations (different underlying trained immunity) without any estimation of the underlying immune status or attempt to make the confounders for the groups being compared to be equal.
- About 8 out of 10 COVID-19 deaths have been in aged >65 years old.
- BCG vaccination is known to provide-cross protection from a number of unrelated diseases.
- BCG vaccination given in childhood protects children from milliary tuberculosis and the conferred protective trained-Immunity correlate or ‘TIC’ (loosely equals ‘tuberculin positivity’) wanes away within 5 years from most in the absence of boosters or rechallenge from environmental *Mycobacterium* spp.

New Findings

- Disregarding BCG vaccination coverage or policy, the prevailing TIC correlate of populations predict protection from COVID-19 in socially similar European countries, *i*.*e*., the countries which are more similar to each other than other parts of the world with regard to various supposed confounders (e.g., exposure, phase of pandemic, health services, social support, food, genetic relatedness etc.).

Recommendations for Policy and Practice

- The planned and ongoing studies or clinical trials assessing the effectiveness of BCG vaccination in protecting populations against COVID-19 or making them vulnerable to COVID-19 should include TIC correlates information of the participants (both controls and vaccinated) to arrive at dependable conclusions about potential benefit of BCG vaccination in controlling COVID-19 infections and mortality.

## INTRODUCTION

There have been efforts to understand and explain the differential impact of COVID-19 on populations in pursuance of identifying protective variables that could predict the impact or be applied for intervention. Studies published in PNAS and Science Advances (Escobar et al. 2020, Berg et al. 2020) had endeavored to explain/model the differential effect on populations based on ‘trained immunity’ correlates of countries’ as per bacillus Calmette–Guérin (BCG) vaccination rates after meticulous correction/fitting of the data for supposed major confounders like age, population density, development status, BCG coverage/implementation using the infections and mortality data from an early stage of pandemic (till April 22, 2020). Later, last year, other communications in PNAS (Lindestam Arlehamn et al 2020, Patella et al. 2020) have failed to find support for the association previously observed between BCG vaccination policy/coverage and the impact of COVID-19 on populations when using updated data set. More recently, a study published by the Citizen science initiative of COVID-BCG Collaborative Working Group in *Transboundary Emerging Diseases* in April 2021 goes on to indicate BCG childhood vaccination as a risk factor for COVID-19 (de la Fuente et al. 2021). These conflicting assertions stem from fundamentally misplaced presumptions that BCG vaccination in childhood would provide lifelong protective or adverse effects completely disregarding the longevity of BCG vaccination conferred immunological correlates that seldomly last > 5 years in the absence of revaccination, rechallenge, or exposure to environmental *Mycobacterial spp*. (Singh 2020a, Menzies 1999, Singh et al. 2020a).

The extrapolation of associative observations made previously (Escobar et al. 2020, Berg et al. 2020) linking BCG vaccination coverage to reduced COVID-19 impact on populations was expected to disappear (Lindestam Arlehamn et al. 2020, Patella et al. 2020, Singh 2020a) as the ‘trained immunity’ conferred by childhood BCG vaccination usually wanes in <5 years (Singh 2020a, Menzies 1999, Singh et al. 2020a). Hence, the premise of protective ‘trained immunity’ from BCG vaccination given in childhood or to children in a population is not supposed to decrease the severity of infection or supposedly provide any protection in currently aged as the BCG conferred ‘trained immunity’ correlates would have waned away long ago (Singh 2020a, Menzies 1999, Singh et al. 2020a). The use of early-stage pandemic data (Escobar et al. 2020, Berg et al. 2020) when the populations were not evenly exposed along with displayed associations’ inherent disconnect with the mechanism proposed behind the observed protective correlation would make such assertions untenable. It was also highlighted by a study from Israel that was published in JAMA last year that found no significant difference in COVID-19 incidence in a group of individuals vaccinated and unvaccinated with BCG in their childhood (Hamiel et al 2020). The same may apply to studies that are using disparate data sets from a later stage of the COVID-19 pandemic and concluding the vaccinated countries to be more protected while overlooking the presence of countries with minimal COVID-19 effect in no BCG vaccination policy countries as well (Islam et al. 2021). Similarly, the studies trying to correlate childhood BCG vaccination to higher COVID-19 incidence or as a risk factor are also potentially indefensible due to gross overlooking of the basic facts about the longevity of trained immunity conferred from BCG vaccination and associated cross-reaction (de la Fuente et al. 2021). The conclusions drawn by the recent report of the Citizen science initiative of COVID-BCG Collaborative Working Group may call for more conservativism and greater scrutiny due to comparison of groups that have disproportionate representation of individuals from disparate underlying *Mycobacterium spp*. conferred background trained immunity as suggested by us previously (Singh et al. 2020a).

We reason, the dependability on the correlative associations as well as conclusions presented in previous studies (Escobar et al. 2020, Berg et al. 2020, Lindestam Arlehamn et al. 2020, Patella et al. 2020, de la Fuente et al. 2021) would have tremendously improved on considerations: a) direct measure of prevailing supposed protective ‘trained immunity’ correlate (TIC) as a result of populations exposure to *Mycobacterium* spp. or BCG vaccination (Singh 2020a, Menzies 1999, Singh et al. 2020a), *i*.*e*., Tuberculin positivity [TIC of BCG given at birth wanes within <5 years (Menzies 1999), so chances of supposed heterologous protection (de la Fuente et al. 2021, Islam et al. 2021, O’Neill et al 2020) of elderly from childhood vaccination are remote]; b) analysis of countries at a similar stage of the pandemic; c) underlying confounders including potential contributory variables (e.g., Vitamin D, Zinc) (Singh et al. 2020b, Singh 2020b, Singh et al. 2021); d) the correlations observed, at any time, to be the total sum of the effects from protective variable and preventive/curative measures in place (e.g., social distancing norms and adherence, medical infrastructure/support).

The European populations with quite dissimilar BCG coverage (including no vaccination) (Zwerling et al 2020) that have experienced differential COVID-19 impact (Worldometer 2020) offer an excellent opportunity to evaluate the alternative hypothesis that the ‘trained immunity from *Mycobacterium* spp. exposure (BCG or environmental NOT the childhood vaccination coverage *per se*) could be protective in COVID-19 as suggested by us previously. It would be theoretically better equipped to predict the outcome or potentially flattened curve if any such association exists, that may have a cause and effect relationship. The current analysis of TIC and COVID-19 data from 24 socially similar European countries completely disregarding their vaccination coverage or policy, support a potential protective role for the prevalent TIC of populations on COVID-19 infections and mortality.

## MATERIAL AND METHODS

The COVID-19 incidence and mortality data for the European countries (Supp. Table 1) was obtained from Worldometer (Worldometer 2020) and that of East and West Germany states from https://www.citypopulation.de/en/germany/covid/ [Accessed on 10 October 2020] and previously published estimates (Singh et al. 2020a). The latent tuberculosis infection (LTBI) (WHO 2018) prevalence estimates (i.e., ‘trained immunity’ correlate) were from Institute for Health Metrics and Evaluation (IHME 2018). All statistical estimations and correlation analysis of the COVID-19 incidence and mortality with TIC or LTBI prevalence of populations (Average, Standard deviation (STDEV), Standard Error (Std. Err.), F-value, Correlation/ Pearson coefficient (r/R), regression, *etc*) were performed using Microsoft Excel 2019. The *p*-values <0.05 were considered significant unless explicitly stated otherwise. The methodology employed has been essentially the same as described previously (Singh et al. 2020a, Singh et al. 2020b, Singh et al. 2021).

## RESULTS & DISCUSSION

Analysis of the updated COVID-19 data (till 28 August, Sup. Table 1, from Worldometer 2020) from 24 European countries with similar confounders (refer Escobar et al. 2020, and additional), stage of the pandemic, without any exclusions (applied in Escobar et al. 2020 and Lindestam Arlehamn et al. 2020) consistently displayed protective/negative correlation with the direct measure of desired heterologous TIC of populations [i.e., tuberculin positivity without active tuberculosis disease; referred by WHO as Latent tuberculosis infection (LTBI) for the management purposes (WHO 2018, IHME 2018)]. Higher LTBI prevalence populations consistently displayed lower COVD-19 incidence/mortality per million (Sup. Table 1). The overall cases and mortality among European countries with similar confounders (Escobar et al. 2020, Berg et al. 2020, Singh et al. 2020a) consistently remained negatively and significantly correlated with the prevalence of trained immunity correlate(%LTBI) for populations (Sup. Fig. 1). The correlative association displayed dependence on the phase of the pandemic (Sup. Fig. 2 &3). The countries with lower LTBI prevalence (<10%) reported higher incidence and fatalities during the study period (12 March 2020 to 26 August 2020) as compared to their higher LTBI prevalence (>10%) neighbors. The correlation between TI estimate (%LTBI) and cases and mortality for COVID-19 consistently remained negative post-peak-of-infections (Cases per million: *r*(24): -0.79 to -0.57; p-value: <0.004; Mortality per million (*r*(24): -0.63 to -0.45; *p*-value: <0.03). The interim case fatality rates (i-CFR) among low (<10%) LTBI prevalence countries remained much higher than that among high LTBI prevalence countries (Fig. 1 A). With the progression of the COVID-19 pandemic the CFR seems to be steadily increasing while incidence rates had been falling (Fig 1A). The correlations observed shown here are supposed to decline further before the disappearance of COVID-19 due to progressive loss of synchronicity of infections/ pandemic-phases (Sup. Fig. 2), lower prevalence of the protective variable (6.06 - 15.95%) (IHME 2018), the differential response of the study population, etc. not necessarily due to the supposed absence of correlation as proposed (Lindestam Arlehamn et al 2020).

**Fig. 1.**
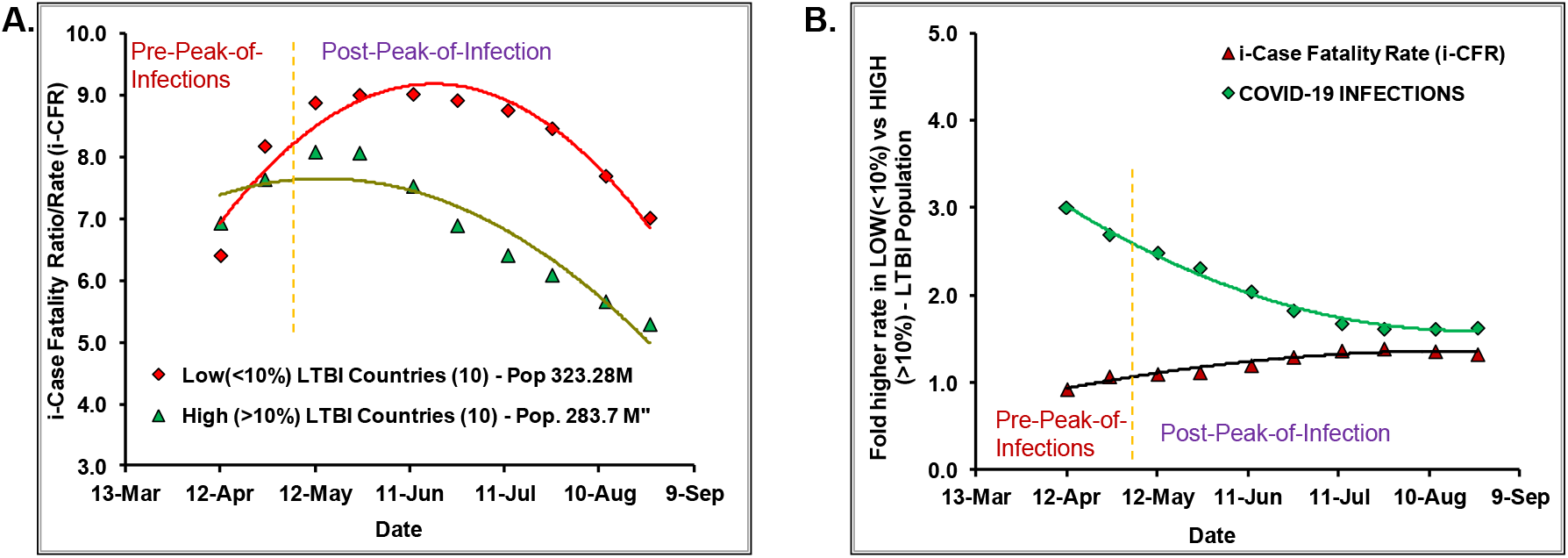
The COVID-19 cases, deaths and CFR consistently remained lower in European populations with higher trained immunity (TI) correlate (>10% LTBI) post-peak-of infections than countries with the lower TI-correlate (<10% LTBI). The TI-correlate indicated a significant consistently negative association with COVID-19 infections [Pearson correlation *r*(24): -0.79 to -0.57, p-value<0.005] and mortality [*r*(24)= -0.63 to - 0.45, p-value<0.05)] for the whole time period (12 March to 26 August 2020). See Sup. Fig 1 in conjunction with Sup. Fig. 2 for detailed correlation analysis and its variation with wave of infections across 24 countries. Refer to Sup. Table 1 for updated COVID-19 cases and deaths data for the 24 countries with supposedly similar confounders and at similar stage of pandemic included in the study. (A) The i-CFR [(deaths/cases)*100] for Low LTBI countries had remained higher than that of high LTBI countries post infections peak. (B) Low LTBI countries have had relatively higher infections/million population (1.63 fold on 26 August) and consistently higher i-CFR (∼30% on 26 August).

The East and the West Germany States that have been proposed in the early stage of the pandemic to be experiencing differential COVID-19 impact (Escobar et al. 2020, Berg et al. 2020) due to differential BCG coverage and policy provide a unique opportunity to test our assertion that actual trained immunity correlates (%LTBI) to be responsible for differential COVID-19 impact. The estimated TIC (LTBI) of East and the West Germany States are 22.5% and 9.2%, respectively (Singh et al. 2020a). The East Germany States with higher TIC have experienced two-fold cases while more than two-fold fewer deaths from COVID per million populations during the study period (Sup. Table 2; https://www.citypopulation.de/en/germany/covid/). The inclusion or exclusion of city-states did not change the supposed overall protective effect on populations. Similarly, the CFR for East and West Germany states remained significantly different for the whole period (Sup. Table 3). The COVID-19 incidence and death rates remained significantly different between East and West Germany States (Fig. 2A &B) both pre and post-peak-of-infections consistent with the potential protective role of TIC prevalence in populations. The CFR rates also remained consistently different during the study period (10 April – 28 August; Fig. 2C) without requiring any correction factors. However, the differential response gap seen for East and West Germany is showing signs of closing as expected for populations slowly reaching towards stable equilibrium with underlying confounders (Fig. 2C).

**Fig 2.**
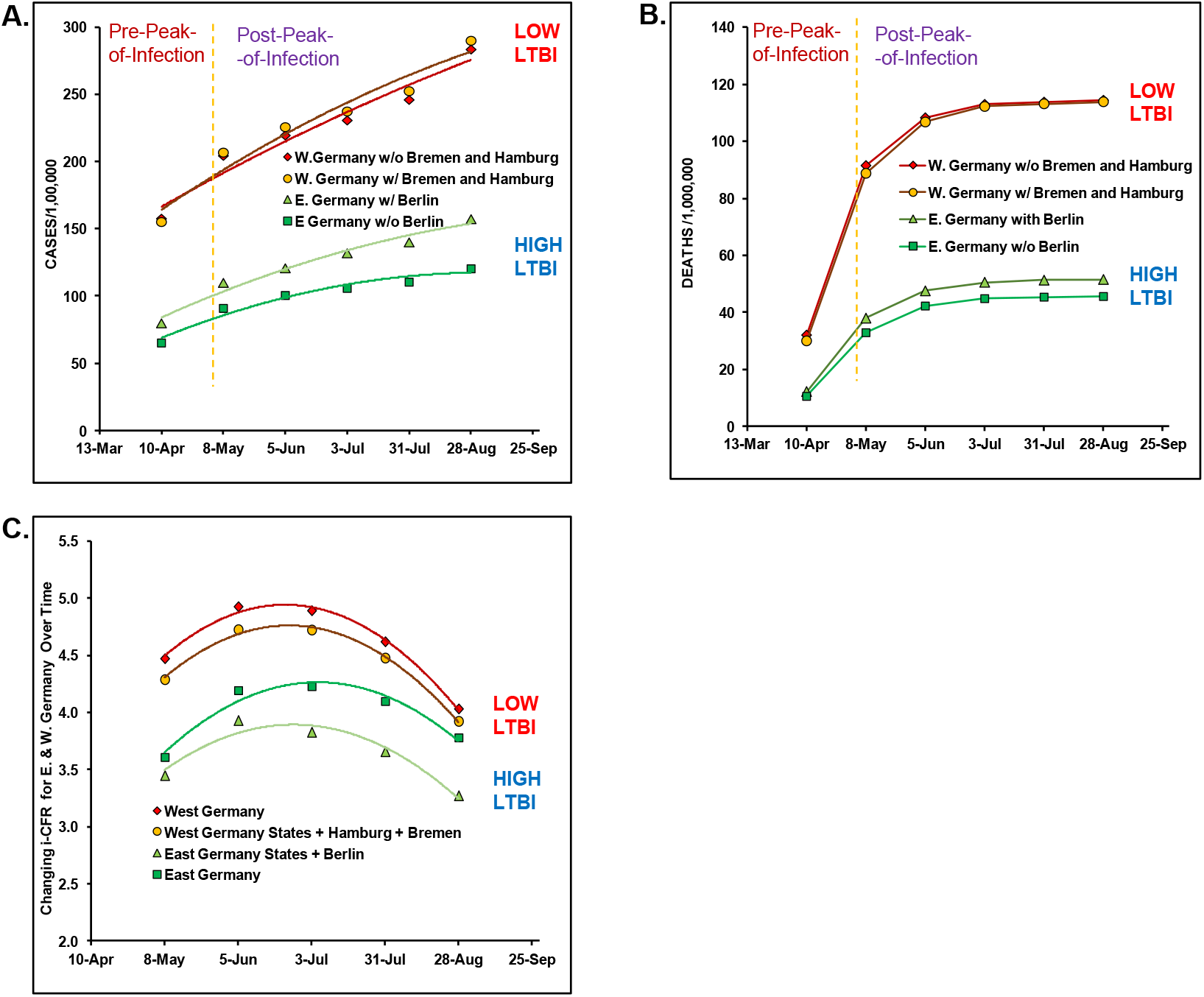
East Germany states with higher trained immunity correlate (%LTBI) as compared to West Germany states (22.5% vs 9.2%) consistently reported lower COVID-19 cases (A), deaths (B) and i-CFR (C) during study period (10 April to 28 August). Refer to Sup. Table 2 for COVID-19 cases and deaths and Sup. Table 3. for i-CFR estimates. E. Germany consistently had 20-30% lower CFR as compared to W. Germany states. The inclusion of Berlin in East Germany region, and that of Hamburg and Bremen in West Germany region decreased the closing trend of the i-CFR with passage of time (compare covariation of Red and Green trend lines with Orange and Light green in (C) possibly indicative of more LTBI positives in Berlin than in Hamburg and Bremen. **In future**, as the pandemic progresses the gap between E. and W. Germany states is expected to close, partially resulting from decrease in the vulnerable population and the concomitant increase in populations overall ‘trained immunity’ as a result of infections.

In conclusion, we believe the incidences, mortality, and i-CFR of COVID-19 would negatively correlate with the trained immunity of populations that have comparable underlying confounders, not the BCG coverage *per se*. To decisively arrive at dependable conclusions about the potential protective benefit of BCG in COVID-19, the ongoing/planned randomized controlled trials (currently total trials listed are 29 as on December 5, 2021) should consciously consider including measures of TIC (Clinical Trials.gov 2021, Hamiel et al. 2020) as - a) all individuals immunized do not respond equally (up to 10-15% could be non-responders), b) small study groups of higher background trained immunity could fail to indicate any protective effect. Currently under development COVID-19 vaccines still have a long way to go and be available in sufficient supply to cover the whole global population at the same time to confer the much-needed and touted ‘herd immunity’ whereas BCG is readily available which can be scaled up at a lower cost to provide the needed respite to vulnerable populations especially in poor countries. Any potential protective effect displayed by BCG vaccination in the ongoing trials, especially in aged and persons with comorbidities who are currently accounting for more than 90% deaths, could help provide hope in the current scenario.

## Data Availability

Included in the manuscript and available at references/links indicated.

## Acknowledgment

The institutional support to the laboratory of SS from the Institute of Eminence (IoE) seed grant, Banaras Hindu University is acknowledged.

## Availability of data and material

*All available in the manuscript and the supplementary information as well as indicated references*.

## Authors’ contributions

SS designed the research, analyzed the data, and wrote the paper; DK and RK contributed to the analysis, drafting and revision.

## Ethics Statement

*‘The authors confirm that the ethical policies of the journal, as noted on the journal’s author guidelines page, have been adhered to. No ethical approval was required with the current design of the study*

---

**Sup. Fig.1.**
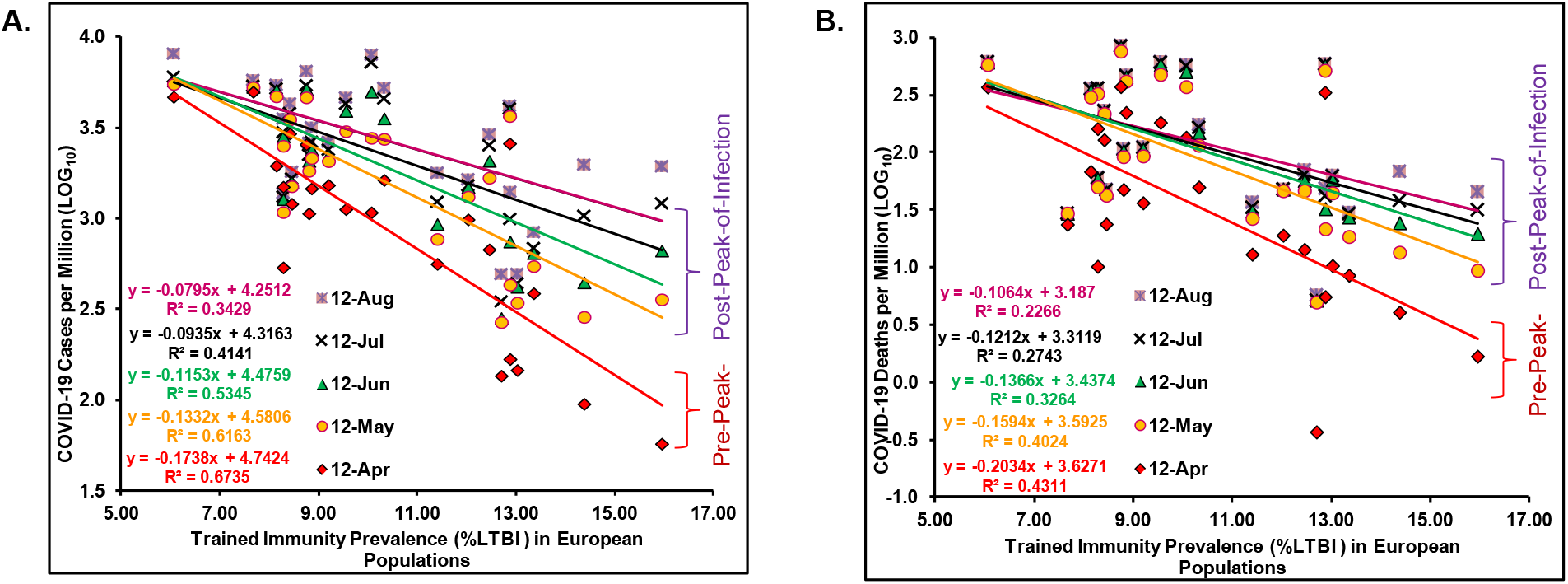
The COVID-19 cases (A) and Deaths (B) in European countries with similar confounders and stage of pandemic consistently remained negatively and significantly correlated with Trained immunity prevalence (est. %LTBI) starting from March 12 to August 26. See Supp. Fig 2 for correlation analysis for the period starting from 12 March to 26 August 2020 covering the duration upto April 22 of ref.1 and August 1 reference point of ref. 2.

**Sup. Fig 2.**
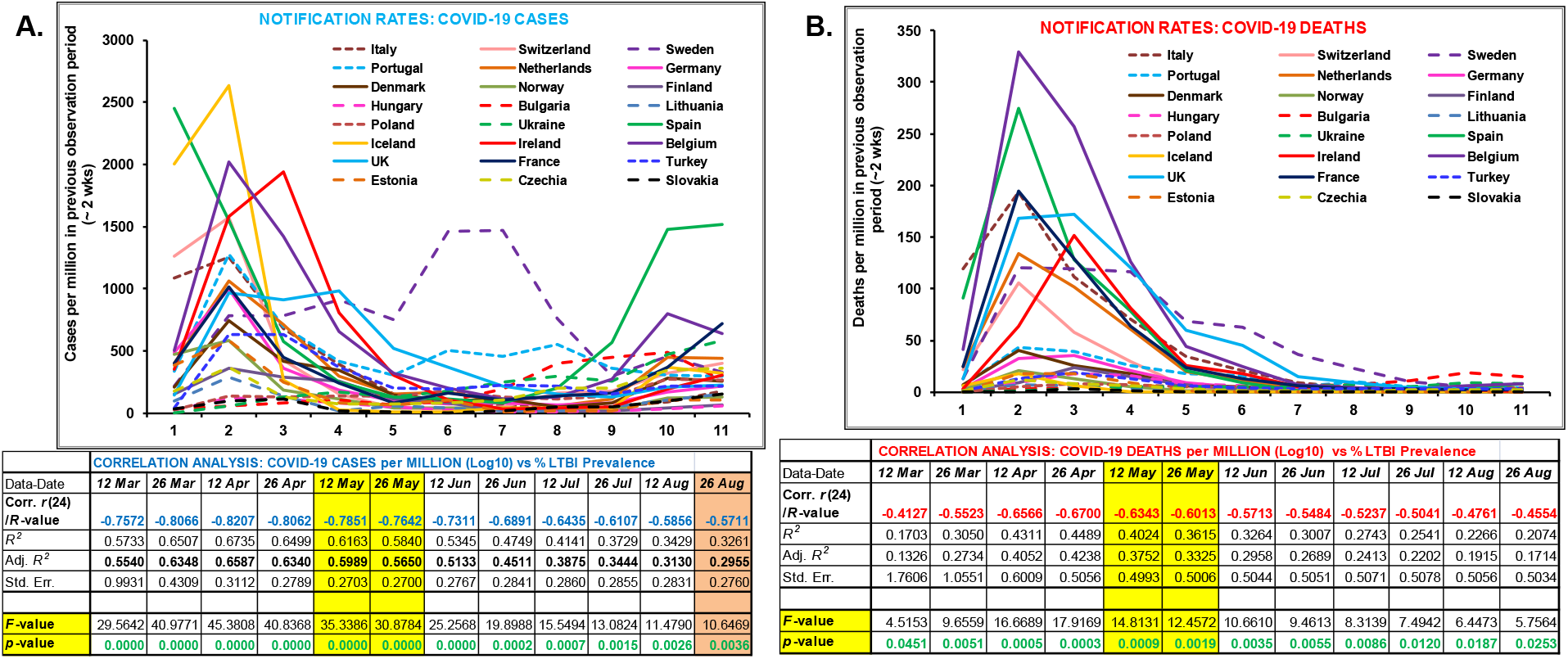
The correlation between underlying prevailing trained immunity correlate (%LTBI) of European populations with COVID-19 cases per million (A) and deaths per million (B) and its dependence on phase of pandemic. The observed correlation (see bottom correlation analysis table) consistently remained negative for the period. The notification rates for countries with >10% LTBI is indicated by broken lines. Correlation remained high with the synchronicity of 1^st^ peak of infections (see corresponding notification rates graph above for the data date indicated in the table below) and been on decline since then partially resulting from the loss of synchroncity, populations response, acquired immunity and understandably and importantly the changing reporting and management practices. ***Refer to Sup. Fig. 3 from European CDC that more accurately reflects the waves of infections/deaths from starting not explicitly observable in the figure one presented here due to coarse methodology employed***. The response of populations had been more stringent and uniform for 1^st^ wave of infections. ***Note:* The highlighted *12-May and 26-May values*** <u>***could*** reflect the assumed total sum of actual maximum achievable correlation for potential ‘trained immunity’ along with current confounders and the stringent measures put in place by the countries to reduce the spread of COVID-19 – ***NOT*** necessarily due to only the prevailing trained immunity of the populations as a result of BCG coverage or implementation alone as assumed (ref.1 and 2.)</u>. <u>**Even if there is a cause and effect relationship**</u>, **the expected protective covariation** (correlation) would expectedly further go down for reasons mentioned above. It may soon reach **0.2-0.4 for the study populations** primarily <u>*due to increasingly heterogenous (loosened) response combined with actual low trained immunity prevalence)*</u>..

**Supplementary Fig 3.**
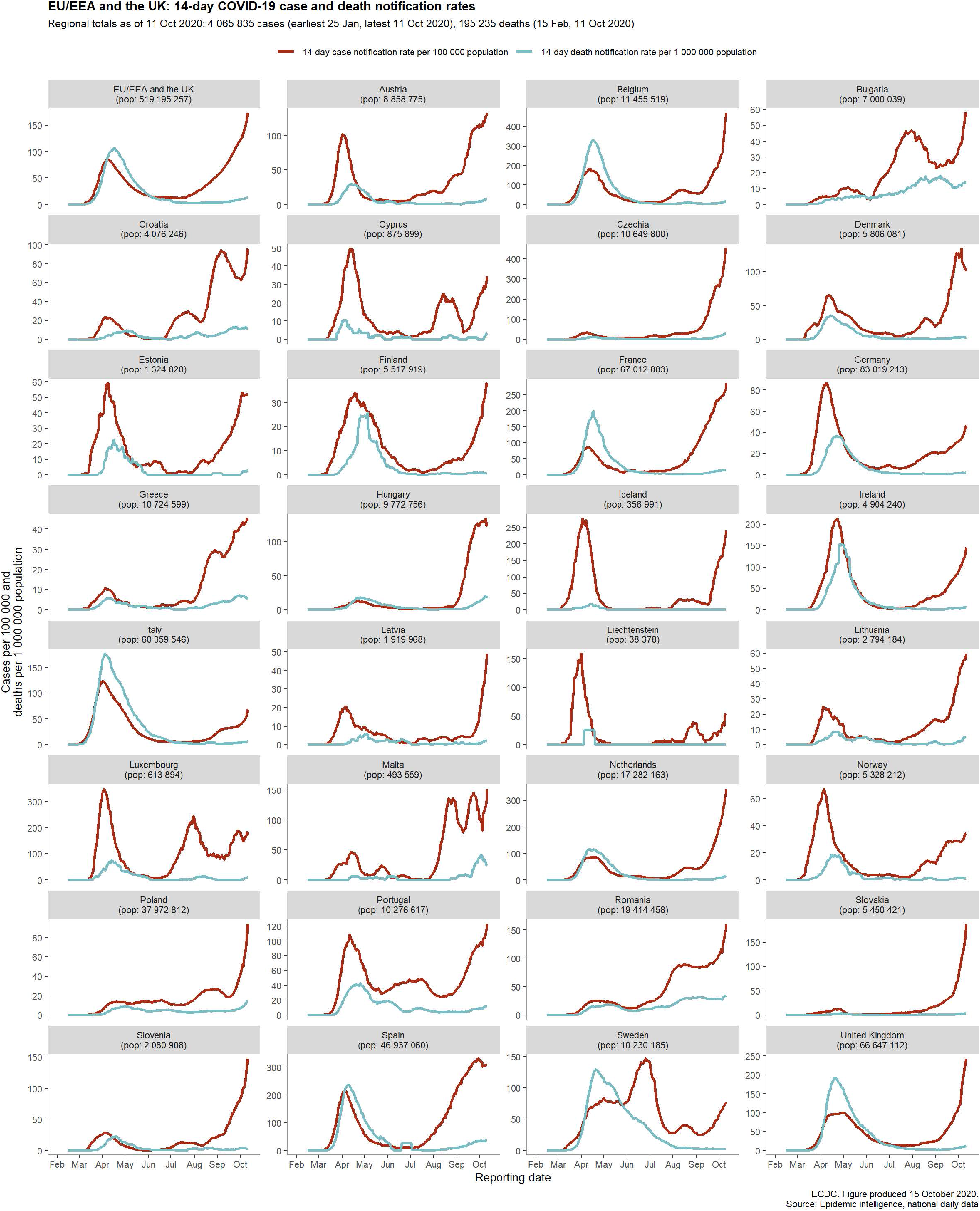

**Sup. Table 1.** COVID-19 data [Cases per million (CpM) and deaths per million DpM]of European countries.

|  | CpM 12 Mar | DpM 12 Mar | CpM 26 Mar | DpM 26 Mar | CpM 12 Apr | DpM 12 Apr | CpM 26 Apr | DpM 26 Apr | CpM 12 May | DpM 12 May | CpM 26 May | DpM 26 May | CpM 12 Jun | DpM 12 Jun | CpM 26 Jun | DpM 26 Jun | CpM 12 Jul | DpM 12 Jul | CpM 26 Jul | DpM 26 Jul | CpM 12 Aug | DpM 12 Aug | CpM 26 Aug | DpM 26 Aug | Population | LTBI 2017 (%) |
| --- | --- | --- | --- | --- | --- | --- | --- | --- | --- | --- | --- | --- | --- | --- | --- | --- | --- | --- | --- | --- | --- | --- | --- | --- | --- | --- |
| Spain | 665.43 | 1.84 | 3118.24 | 93.35 | 4667.48 | 368.05 | 5243.71 | 495.96 | 5493.08 | 574.36 | 5622.58 | 595.47 | 5738.28 | 604.71 | 5819.68 | 606.06 | 6019.13 | 607.49 | 6584.73 | 608.11 | 8063.46 | 611.21 | 9581.54 | 619.60 | 46757733 | 6.06 |
| Iceland | 342.51 | 0.00 | 2347.79 | 5.85 | 4979.54 | 23.42 | 5245.93 | 29.27 | 5272.28 | 29.27 | 5281.06 | 29.27 | 5289.84 | 29.27 | 5327.90 | 29.27 | 5365.96 | 29.27 | 5406.94 | 29.27 | 5772.87 | 29.27 | 6094.88 | 29.27 | 341598 | 7.67 |
| Ireland | 14.15 | 0.20 | 367.73 | 3.84 | 1950.46 | 67.52 | 3892.64 | 218.94 | 4697.25 | 300.01 | 4999.08 | 325.68 | 5103.19 | 343.88 | 5136.34 | 348.93 | 5181.02 | 352.98 | 5232.17 | 356.21 | 5425.64 | 358.64 | 5733.94 | 359.24 | 4946514 | 8.14 |
| Finland | 19.67 | 0.00 | 172.86 | 0.90 | 536.62 | 10.10 | 825.68 | 34.28 | 1083.16 | 49.62 | 1195.93 | 56.30 | 1276.23 | 58.64 | 1297.52 | 59.18 | 1315.92 | 59.18 | 1333.97 | 59.36 | 1378.89 | 60.09 | 1443.85 | 60.45 | 5542120 | 8.28 |
| Netherlands | 35.82 | 0.29 | 433.52 | 25.32 | 1492.73 | 159.68 | 2207.86 | 261.07 | 2507.67 | 321.45 | 2659.00 | 341.64 | 2827.19 | 353.13 | 2917.27 | 356.05 | 2976.60 | 357.91 | 3088.85 | 358.20 | 3536.95 | 359.43 | 3973.74 | 362.58 | 17141028 | 8.29 |
| Switzerland | 100.18 | 0.81 | 1363.11 | 22.16 | 2933.15 | 127.64 | 3353.93 | 185.81 | 3506.16 | 215.47 | 3550.13 | 221.01 | 3584.98 | 223.66 | 3633.80 | 226.43 | 3795.03 | 227.13 | 3971.49 | 228.17 | 4289.68 | 229.78 | 4690.84 | 231.17 | 8664751 | 8.42 |
| Norway | 147.38 | 0.18 | 621.22 | 2.58 | 1202.10 | 23.58 | 1386.69 | 37.03 | 1502.76 | 42.00 | 1544.40 | 43.29 | 1588.06 | 44.58 | 1627.11 | 45.87 | 1654.56 | 46.43 | 1679.62 | 46.98 | 1802.32 | 47.16 | 1935.15 | 48.64 | 5428015 | 8.46 |
| Belgium | 34.40 | 0.34 | 537.60 | 41.30 | 2556.27 | 370.16 | 3977.84 | 627.10 | 4637.01 | 753.16 | 4953.97 | 797.05 | 5157.80 | 821.97 | 5268.77 | 829.30 | 5398.11 | 833.52 | 5667.21 | 836.63 | 6467.45 | 843.18 | 7108.87 | 851.72 | 11597764 | 8.75 |
| Denmark | 116.30 | 0.00 | 323.87 | 7.07 | 1065.31 | 47.11 | 1479.60 | 72.82 | 1827.45 | 90.93 | 1971.87 | 97.14 | 2072.12 | 102.49 | 2187.04 | 104.22 | 2233.80 | 105.08 | 2318.69 | 105.77 | 2600.29 | 107.15 | 2853.42 | 107.50 | 5795502 | 8.81 |
| France | 44.04 | 0.93 | 446.50 | 25.97 | 1461.06 | 220.22 | 1907.82 | 349.69 | 2147.53 | 412.99 | 2229.12 | 436.54 | 2393.48 | 449.47 | 2495.31 | 455.66 | 2635.48 | 459.23 | 2795.92 | 462.50 | 3165.48 | 465.12 | 3883.60 | 467.77 | 65296963 | 8.86 |
| Germany | 32.75 | 0.07 | 524.15 | 3.19 | 1525.21 | 36.05 | 1882.08 | 71.29 | 2065.81 | 92.31 | 2162.64 | 101.38 | 2233.77 | 105.73 | 2319.04 | 107.67 | 2385.26 | 108.96 | 2466.27 | 109.79 | 2634.58 | 110.66 | 2851.10 | 111.56 | 83827321 | 9.20 |
| UK | 7.89 | 0.13 | 155.54 | 12.92 | 1123.82 | 180.71 | 2038.02 | 352.76 | 3019.69 | 472.95 | 3536.57 | 532.41 | 3906.24 | 577.36 | 4125.05 | 592.38 | 4274.81 | 601.16 | 4406.99 | 605.08 | 4618.52 | 608.29 | 4840.00 | 610.29 | 67943420 | 9.55 |
| Sweden | 76.27 | 0.10 | 296.36 | 13.26 | 1079.30 | 134.03 | 1865.89 | 253.13 | 2775.15 | 369.85 | 3523.17 | 438.21 | 4982.21 | 500.53 | 6450.65 | 536.53 | 7209.05 | 558.89 | 7504.02 | 568.68 | 7961.82 | 573.53 | 8291.71 | 575.31 | 10109375 | 10.07 |
| Portugal | 7.65 | 0.00 | 347.73 | 5.89 | 1627.29 | 49.45 | 2324.22 | 88.60 | 2738.77 | 114.11 | 3042.34 | 131.67 | 3549.91 | 147.67 | 4009.69 | 152.57 | 4563.66 | 162.88 | 4921.99 | 168.47 | 5223.11 | 173.08 | 5521.49 | 177.30 | 10191814 | 10.33 |
| Czechia | 10.92 | 0.00 | 189.32 | 0.84 | 561.32 | 12.88 | 693.23 | 20.54 | 769.50 | 26.42 | 846.98 | 29.59 | 929.87 | 30.71 | 1032.56 | 32.39 | 1231.77 | 32.86 | 1432.48 | 34.63 | 1780.68 | 36.50 | 2142.51 | 39.02 | 10712210 | 11.41 |
| Estonia | 20.35 | 0.00 | 405.52 | 0.75 | 986.67 | 18.84 | 1238.43 | 36.93 | 1316.07 | 45.98 | 1382.40 | 47.49 | 1484.91 | 47.49 | 1496.97 | 47.49 | 1518.08 | 47.49 | 1533.15 | 47.49 | 1638.68 | 47.49 | 1741.94 | 48.24 | 1326680 | 12.03 |
| Turkey | 0.01 | 0.00 | 42.96 | 0.89 | 674.17 | 14.18 | 1303.58 | 33.20 | 1674.60 | 46.09 | 1879.22 | 52.05 | 2074.01 | 56.56 | 2302.37 | 59.95 | 2521.14 | 63.48 | 2676.28 | 66.44 | 2892.80 | 69.73 | 3107.22 | 73.19 | 84482851 | 12.46 |
| Slovakia | 3.85 | 0.00 | 41.39 | 0.00 | 135.90 | 0.37 | 252.56 | 3.30 | 268.31 | 4.94 | 277.10 | 5.13 | 282.41 | 5.13 | 300.91 | 5.13 | 348.16 | 5.13 | 399.08 | 5.13 | 492.67 | 5.68 | 647.61 | 6.04 | 5460073 | 12.70 |
| Italy | 250.17 | 16.87 | 1333.08 | 136.33 | 2586.60 | 330.19 | 3269.98 | 441.89 | 3659.54 | 512.71 | 3814.12 | 546.69 | 3909.39 | 567.70 | 3970.11 | 576.73 | 4021.39 | 580.80 | 4071.91 | 583.34 | 4164.18 | 585.29 | 4343.26 | 586.59 | 60447245 | 12.87 |
| Poland | 0.819 | 0 | 27.77 | 0.37 | 167.941 | 5.496 | 297.86 | 13.845 | 431.373 | 21.429 | 571.544 | 26.607 | 745.139 | 32.103 | 875.085 | 37.308 | 991.397 | 41.43 | 1126.178 | 43.967 | 1399.359 | 48.115 | 1666.543 | 52.237 | 37850425 | 12.88015 |
| Hungary | 1.66 | 0.00 | 27.03 | 1.04 | 146.02 | 10.25 | 258.90 | 28.17 | 343.09 | 44.01 | 390.52 | 51.68 | 419.73 | 57.48 | 427.39 | 59.86 | 438.47 | 61.62 | 459.28 | 61.72 | 493.77 | 62.65 | 547.62 | 63.59 | 9656316 | 13.03 |
| Lithuania | 1.102 | 0 | 100.651 | 1.469 | 386.807 | 8.449 | 523.824 | 15.061 | 545.496 | 18.367 | 600.597 | 23.142 | 643.576 | 27.183 | 663.412 | 28.652 | 685.085 | 29.02 | 735.043 | 29.387 | 838.632 | 29.754 | 989.608 | 31.224 | 2725537 | 13.35834 |
| Bulgaria | 1.007 | 0.144 | 34.828 | 0.432 | 95.129 | 4.03 | 179.465 | 7.915 | 286.395 | 13.384 | 350.15 | 18.709 | 444.128 | 24.178 | 634.387 | 30.367 | 1032.605 | 37.706 | 1484.073 | 48.644 | 1974.83 | 67.785 | 2243.524 | 82.321 | 6953085 | 14.37964 |
| Ukraine | 0.00 | 0.00 | 2.584 | 0.091 | 57.416 | 1.669 | 185.783 | 4.596 | 357.801 | 9.329 | 485.78 | 14.245 | 664.704 | 19.527 | 914.808 | 24.398 | 1208.289 | 31.372 | 1461.777 | 36.356 | 1933.243 | 45.045 | 2517.163 | 53.003 | 43757273 | 15.95401 |
| Average (for countries <10 % LTBI) | 130.04 | 0.40 | 867.68 | 20.37 | 2124.48 | 136.19 | 2786.82 | 228.00 | 3146.65 | 279.55 | 3308.86 | 298.10 | 3430.93 | 309.58 | 3512.90 | 313.42 | 3602.97 | 315.70 | 3746.07 | 317.17 | 4146.34 | 319.16 | 4582.58 | 321.65 |  | 8.37 |
| STDEV | 192.73 | 0.55 | 938.59 | 26.17 | 1420.07 | 128.68 | 1509.03 | 196.92 | 1539.49 | 236.51 | 1572.01 | 251.32 | 1583.06 | 260.70 | 1588.63 | 263.46 | 1619.76 | 265.23 | 1713.63 | 266.23 | 2009.51 | 267.91 | 2311.44 | 270.34 |  | 0.88 |
|  |  |  |  |  |  |  |  |  |  |  |  |  |  |  |  |  |  |  |  |  |  |  |  | Pop. | 323282729 |  |
| Average (for countries >10 % LTBI) | 31.15 | 1.43 | 237.43 | 13.45 | 708.71 | 49.15 | 1032.81 | 78.93 | 1263.84 | 102.22 | 1430.33 | 115.43 | 1677.50 | 126.35 | 1923.19 | 132.62 | 2147.42 | 137.72 | 2317.11 | 141.19 | 2566.15 | 145.39 | 2813.35 | 149.01 |  | 12.62 |
| STDEV | 72.22 | 4.87 | 372.47 | 38.89 | 765.70 | 96.01 | 1002.28 | 133.70 | 1184.71 | 163.84 | 1317.75 | 180.57 | 1600.43 | 194.35 | 1916.26 | 201.58 | 2091.14 | 205.55 | 2148.41 | 206.93 | 2215.65 | 206.71 | 2261.31 | 205.97 |  | 1.61 |
|  |  |  |  |  |  |  |  |  |  |  |  |  |  |  |  |  |  |  |  |  |  |  |  | Pop. | 283672884 |  |
| 1. Note: Countries name in Black from ref 1.) |  |  |  |  |  |  |  |  |  |  |  |  |  |  |  |  |  |  |  |  |  |  |  |  |  |  |
| 2. CpM: Cases per million; DpM: Deaths per million; Mar: March; Apr: April; Jun: June; Jul: July; Aug: August 2020 |  |  |  |  |  |  |  |  |  |  |  |  |  |  |  |  |  |  |  |  |  |  |  |  |  |  |
| 3. LTBI estimates from ref 8. Global Burden of Disease Collaborative Network. Global Burden of Disease Study 2017 (GBD 2017) Results. Seattle, United States: Institute for Health Metrics and Evaluation (IHME), 2018. |  |  |  |  |  |  |  |  |  |  |  |  |  |  |  |  |  |  |  |  |  |  |  |  |  |  |
| [Available from http://ghdx.healthdata.org/gbd-results-tool; search term combination "Prevalence - Latent Tuberculosis Infection - Sex: Both - Age: All Ages (Percent)"] |  |  |  |  |  |  |  |  |  |  |  |  |  |  |  |  |  |  |  |  |  |  |  |  |  |  |
| 4. STDEV: Standard deviation |  |  |  |  |  |  |  |  |  |  |  |  |  |  |  |  |  |  |  |  |  |  |  |  |  |  |

**Sup. Table 2.** COVID-19 Cases and Deaths of erstwhile East and West Germany states (Ref.1)

| Table 2. COVID-19 Cases and Deaths of erstwhile East and West Germany states (Ref.1) |  |  |  |  |  |  |  |  |  |  |  |  |  |
| --- | --- | --- | --- | --- | --- | --- | --- | --- | --- | --- | --- | --- | --- |
| **EAST GERMANY (Region)** |  | COVID-19 Cases per 100k population |  |  |  |  |  | COVID-19 Deaths per million population |  |  |  |  |  |
| States\Dates | 10-Apr | 8-May | 5-Jun | 3-Jul | 31-Jul | 28-Aug |  | 10-Apr | 8-May | 5-Jun | 3-Jul | 31-Jul | 28-Aug |
| *Berlin: City State* | 133.5 | 173.4 | 193.5 | 232.2 | 256.6 | 308.3 |  | NA | 44.42 | 54.5 | 58.32 | 60.23 | 60.77 |
| Brandenburg | 84.74 | 122.6 | 129.2 | 136.2 | 142 | 154.2 |  | 19.03 | 51.95 | 61.86 | 65.82 | 66.22 | 66.22 |
| Mecklenburg-Vorpommern<br>[Mecklenburg-Western<br>Pomerania] | 38.31 | 45.46 | 48.13 | 50 | 55.09 | 62.93 |  | 6.84 | 11.81 | 12.44 | 12.44 | 12.44 | 12.44 |
| Sachsen [Saxony] | 94.33 | 121.2 | 130.8 | 134.1 | 136.4 | 147.2 |  | 15.72 | 45.19 | 52.06 | 55.01 | 55.26 | 55.26 |
| Sachsen-Anhalt [Saxony-Anhalt] | 54.9 | 75.09 | 78.55 | 86.11 | 92.17 | 102.1 |  | 8.201 | 21.87 | 25.06 | 26.88 | 28.25 | 29.16 |
| Thüringen [Thuringia] | 73.55 | 122.3 | 145 | 153.5 | 158.6 | 169.7 |  | 11.25 | 52.5 | 79.22 | 84.84 | 85.31 | 85.31 |
| **Average ( without Berlin)** | **65.27** | **91.01** | **100.62** | **105.93** | **110.57** | **120.48** |  | **10.50** | **32.84** | **42.20** | **44.79** | **45.32** | **45.54** |
| **Average (with Berlin)** | **79.89** | **110.01** | **120.86** | **132.02** | **140.14** | **157.41** |  | **12.21** | **37.96** | **47.52** | **50.55** | **51.29** | **51.53** |
| **WEST GERMANY (Region)** |  | COVID-19 Cases per 100k population |  |  |  |  |  | COVID-19 Deaths per million population |  |  |  |  |  |
| *Bremen: City State* | 72.52 | 164.1 | 225.6 | 246.9 | 262.8 | 292 |  | 19.08 | 46.98 | 64.59 | 77.8 | 80.74 | 82.21 |
| *Hamburg: City State* | 218.2 | 269 | 275.7 | 281.8 | 294.5 | 340.6 |  | 28.69 | 108.8 | 137 | 140.7 | 140.7 | 141.3 |
| Baden-Württemberg | 235.7 | 301.5 | 315.5 | 323.2 | 337.6 | 378.5 |  | 55.31 | 136.5 | 160.2 | 165.7 | 165.7 | 166.4 |
| Bayern [Bavaria] | 264.7 | 340.3 | 361.5 | 371.7 | 389.7 | 437.2 |  | 54.93 | 161.1 | 189 | 197.9 | 199.3 | 199.8 |
| Hessen [Hesse] | 101.8 | 143.4 | 162.6 | 174.6 | 193.7 | 246.9 |  | 17.81 | 64.09 | 76.65 | 80.95 | 81.74 | 82.7 |
| Niedersachsen [Lower<br>Saxony] | 102.8 | 136.7 | 156.7 | 171 | 182.1 | 209.8 |  | 21.39 | 61.67 | 75.56 | 79.19 | 80.56 | 81.44 |
| Nordrhein-Westfalen [North<br>Rhine-Westphalia] | 150.1 | 195.8 | 215.7 | 245.8 | 275.7 | 327.8 |  | 27.97 | 77.84 | 90.1 | 93.94 | 95.34 | 96.56 |
| Rheinland-Pfalz [Rhineland-Palatinate] | 121.1 | 154.3 | 165.6 | 173 | 184.9 | 219.7 |  | 14.66 | 46.9 | 56.18 | 57.4 | 58.14 | 58.38 |
| Saarland | 209.2 | 263.4 | 272.9 | 277.6 | 285.7 | 309.7 |  | NA | 141.9 | 168.2 | 176.3 | 176.3 | 176.3 |
| Schleswig-Holstein | 76.76 | 101 | 107 | 110 | 119.9 | 139.6 |  | NA | 42.01 | 50.28 | 52.35 | 53.03 | 53.72 |
| **Average (without Bremen**<br>**& Hamburg)** | **157.77** | **204.55** | **219.69** | **230.86** | **246.16** | **283.65** |  | **32.01** | **91.50** | **108.27** | **112.97** | **113.76** | **114.41** |
| **Average (with Bremen &**<br>**Hamburg)** | **155.29** | **206.95** | **225.88** | **237.56** | **252.66** | **290.18** |  | **29.98** | **88.78** | **106.78** | **112.22** | **113.16** | **113.88** |

Reference 1. https://www.citypopulation.de/en/germany/covid/ [Accessed on 10 October 2020].

**Sup. Table 3.** i-CFR rates in East and West Germany region/ states.

| **Table 3. i-CFR rates in East and West Germany region/ states** |  |  |  |  |  |
| --- | --- | --- | --- | --- | --- |
| **EAST GERMANY (Region)** | **i-CFR RATES AT INDICATED DATES** |  |  |  |  |
| **States\Date** | **8-May** | **5-Jun** | **3-Jul** | **31-Jul** | **28-Aug** |
| *Berlin: City State* | 2.562 | 2.817 | 2.512 | 2.347 | 1.971 |
| Brandenburg | 4.237 | 4.788 | 4.833 | 4.663 | 4.294 |
| Mecklenburg-Vorpommern [Mecklenburg-Western Pomerania] | 2.598 | 2.585 | 2.488 | 2.258 | 1.977 |
| Sachsen [Saxony] | 3.729 | 3.980 | 4.102 | 4.051 | 3.754 |
| Sachsen-Anhalt [Saxony-Anhalt] | 2.913 | 3.190 | 3.122 | 3.065 | 2.856 |
| Thüringen [Thuringia] | 4.293 | 5.463 | 5.527 | 5.379 | 5.027 |
| **Average (w/o Berlin)** | 3.609 | 4.194 | 4.229 | 4.098 | 3.780 |
| **Average (with Berlin)** | 3.450 | 3.932 | 3.829 | 3.659 | 3.274 |
| **WEST GERMANY** |  |  |  |  |  |
| *Bremen: City State* | 2.863 | 2.863 | 3.151 | 3.072 | 2.815 |
| *Hamburg: City State* | 4.045 | 4.969 | 4.993 | 4.778 | 4.149 |
| Baden-Württemberg | 4.527 | 5.078 | 5.127 | 4.908 | 4.396 |
| Bayern [Bavaria] | 4.734 | 5.228 | 5.324 | 5.114 | 4.570 |
| Hessen [Hesse] | 4.469 | 4.714 | 4.636 | 4.220 | 3.350 |
| Niedersachsen [Lower Saxony] | 4.511 | 4.822 | 4.631 | 4.424 | 3.882 |
| Nordrhein-Westfalen [North Rhine-Westphalia] | 3.975 | 4.177 | 3.822 | 3.458 | 2.946 |
| Rheinland-Pfalz [Rhineland-Palatinate] | 3.040 | 3.393 | 3.318 | 3.144 | 2.657 |
| Saarland | 5.387 | 6.163 | 6.351 | 6.171 | 5.693 |
| Schleswig-Holstein | 4.159 | 4.699 | 4.759 | 4.423 | 3.848 |
| **Average (w/o Bremen & Hamburg)** | 4.473 | 4.928 | 4.893 | 4.621 | 4.034 |
| **Average (w/ Bremen & Hamburg)** | 4.290 | 4.727 | 4.724 | 4.479 | 3.924 |

**Note:** East Germany states (est. 22.5% LTBI of vulnerable population (Ref..2) consistently reported lower i-CFR as compared to West Germany States (est. 9.2% LTBI of vulnerable population). E. Germany with higher trained immunity correlate consistently had 20-30% lower CFR as compared to W. Germany states. The inclusion of Berlin in East Germany region, and that of Hamburg and Bremen in West Germany region decreased the closing trend of the i-CFR with passage of time (Compare covariation of Red and Green trend lines with Orange and Light green in Fig. 2C) that could be reflective of more LTBI positives in Berlin as compared to Hamburg and Bremen.

